# Addition of losartan to FOLFORINOX and chemoradiation downregulates pro-invasion and immunosuppression-associated genes in locally advanced pancreatic cancer

**DOI:** 10.1101/2022.06.09.22275912

**Authors:** Yves Boucher, Jessica M. Posada, Sonu Subudhi, Spencer R. Rosario, Liqun Gu, Ashwin S. Kumar, Heena Kumra, Mari Mino-Kenudson, Nilesh P. Talele, Dan G. Duda, Dai Fukumura, Jennifer Y. Wo, Jeffrey W. Clark, David P. Ryan, Carlos Fernandez-Del Castillo, Theodore S. Hong, Mikael J. Pittet, Rakesh K. Jain

## Abstract

**Purpose:** Adding losartan to FOLFIRINOX (FFX) chemotherapy followed by chemoradiation (CRT) resulted in 61% R0 surgical resection in our phase II trial in patients with locally advanced pancreatic cancer (LAPC). Here we identify potential mechanisms of benefit by assessing the effects of neoadjuvant losartan+FFX+CRT versus FFX+CRT on the stromal tumor microenvironment.

**Experimental Design:** We performed a gene expression analysis of RNA extracted from pancreatic cancer tissue sections and immunofluorescence for cancer cells and immune cells using archived surgical samples from patients treated with losartan+FFX+CRT (<u>NCT01591733</u>), FFX+CRT (<u>NCT01591733</u>) or surgery upfront, without any neoadjuvant therapy. We then assessed whether certain gene sets could stratify the overall survival (OS) of patients.

**Results:** Neoadjuvant losartan+FFX+CRT and FFX+CRT increased the expression of genes linked to vascular normalization, transendothelial migration of leukocytes, T cell activation and cytolytic activity, and dendritic cell (DC) related genes versus no neoadjuvant treatment. In comparison to FFX+CRT, losartan+FFX+CRT downregulated pro-invasion, immunosuppression, and M2 macrophages related genes, and upregulated genes associated with tumor suppression, including the p53 pathway. Furthermore, immunostaining revealed significantly less residual disease in lesions treated with losartan+FFX+CRT versus FFX+CRT. Losartan+FFX+CRT also reduced CD4^+^FOXP3^+^ regulatory T cells in PDAC lesions with a complete/near complete response. OS was associated with DC and antigen presentation genes for patients treated with FFX+CRT, and with immunosuppression and invasion genes or DC- and blood vessel-related genes for those treated with losartan+FFX+CRT.

**Conclusions:** Adding losartan to FFX+CRT reduced pro-invasion and immunosuppression related genes, which were associated with improved treatment outcomes in patients with LAPC.

## Introduction

The poor survival of patients with locally advanced and metastatic pancreatic ductal adenocarcinoma (PDAC) is due to its aggressive biology, limited effectiveness of cytotoxic agents, and an immuno-suppressive tumor microenvironment (TME) [1, 2]. The abnormal PDAC TME promotes the recruitment of M2 macrophages, myeloid-derived suppressor cells (MDSCs), neutrophils, and regulatory T cells (Tregs), which secrete inflammatory cytokines (e.g. IL-10, GM-CSF, TGF-□) that can promote PDAC progression and metastasis [3, 4], and impair the infiltration and activity of T cells, NK cells and dendritic cells (DCs) [4-7].

We have shown that angiotensin system inhibitors (ASIs), including the angiotensin receptor blocker losartan, can enhance the delivery and efficacy of cytotoxic agents in (PDAC) models [8]. The mechanisms underlying this benefit include “normalization” of cancer-associated fibroblasts (CAFs) and extracellular matrix (ECM), resulting in blood vessel decompression, improved perfusion, and decreased hypoxia [8, 9]. Based on our experimental results with losartan, we initiated a clinical trial at Massachusetts General Hospital to determine whether losartan improves the effectiveness of the drug cocktail FOLFIRINOX (FFX) and chemoradiation (CRT) in patients with locally advanced PDAC (LAPC). The results of this trial indicated that adding losartan to FFX+CRT was associated with high rates of surgical resection (69.4%) and R0 resection (61%) in LAPC [10].

In a retrospective analysis, we examined the effect of long-term ASI use on the survival of patients with PDAC and explored its potential mechanisms [11]. Our findings indicated that chronic ASI use is independently associated with longer overall survival in non-metastatic PDAC patients [11]. Unbiased gene expression analysis suggested that the improved survival associated with ASI (lisinopril) therapy could be due to normalization of the extracellular matrix, improved oxidative phosphorylation, inhibition of tumor progression (down-regulation of cell cycle, NOTCH, and WNT pathways) and enhanced anti-tumor immunity [11]. ASIs are known to alleviate hypoxia – which suppresses DC activation [12] – and inhibit the activity of tumor-promoting macrophage phenotypes in other cancer types [13, 14]. Cytotoxic agents can also provide antitumor benefit by reprogramming immune cells, including CD8^+^ T cells, DCs, tumor associated macrophages (TAMs) and myeloid derived suppressor cells (MDSCs) [15, 16]. Recent studies have shown that neoadjuvant FFX can increase effector T cells and decrease immunosuppressive cells in patients with PDAC. Neoadjuvant FFX increased the intratumoral recruitment of CD4^+^ T cells, CD8^+^ T cells and MHC-1 expression, and reduced the infiltration of FOXP3^+^ Tregs and CD163^+^ TAMs in LAPC [17]. A recent study using CyTOF revealed that FFX reduced inflammatory monocytes and Tregs, increased Th1 cells and decreased Th2 cells in the peripheral circulation of PDAC patients [18]. Therefore, we expect that losartan and cytotoxics (particularly when used in combination) will reprogram the immunosuppressive human PDAC TME to an immunostimulatory milieu. To test this hypothesis, we obtained resected PDAC samples from patients treated with neoadjuvant FFX+CRT without losartan or losartan+FFX+CRT from two different phase II trials (<u>NCT01591733 and NCT01821729)</u>, and from patients that underwent surgery upfront, without any neoadjuvant therapy. To identify the potential mechanisms of benefit of adding losartan to FFX+CRT, we performed a transcriptome analysis of RNA extracted from surgical specimens. We also used immunofluorescence to assess the infiltration by immune cells in the same PDAC tissues.

## Materials and Methods

### Patient Population and Treatment Plan

PDAC patients enrolled in two completed trials at Massachusetts General Hospital were included in our investigations of the immune and stromal TME in resected PDAC lesions: “Phase II study of preoperative FOLFIRINOX followed by accelerated short course radiation therapy with capecitabine for borderline-resectable pancreatic cancer (<u>NCT01591733</u>),” and “Phase II study of FOLFIRINOX-losartan followed by accelerated short course radiation therapy with capecitabine for locally advanced pancreatic cancer (<u>NCT01821729</u>).” PDAC tissue was obtained from 19 patients with locally advanced PDAC treated with losartan+FFX+CRT, and 20 patients with borderline resectable PDAC treated with FFX+CRT **(Fig. 1A)**. To control for the effects of cytotoxic therapy or losartan, we also included in our study resected PDAC samples (N=9) from patients with stage I or II resectable PDAC who did not receive any neoadjuvant therapy, as a control. This study was approved by the Massachusetts General Hospital Institutional Review Board.

**Figure 1.**
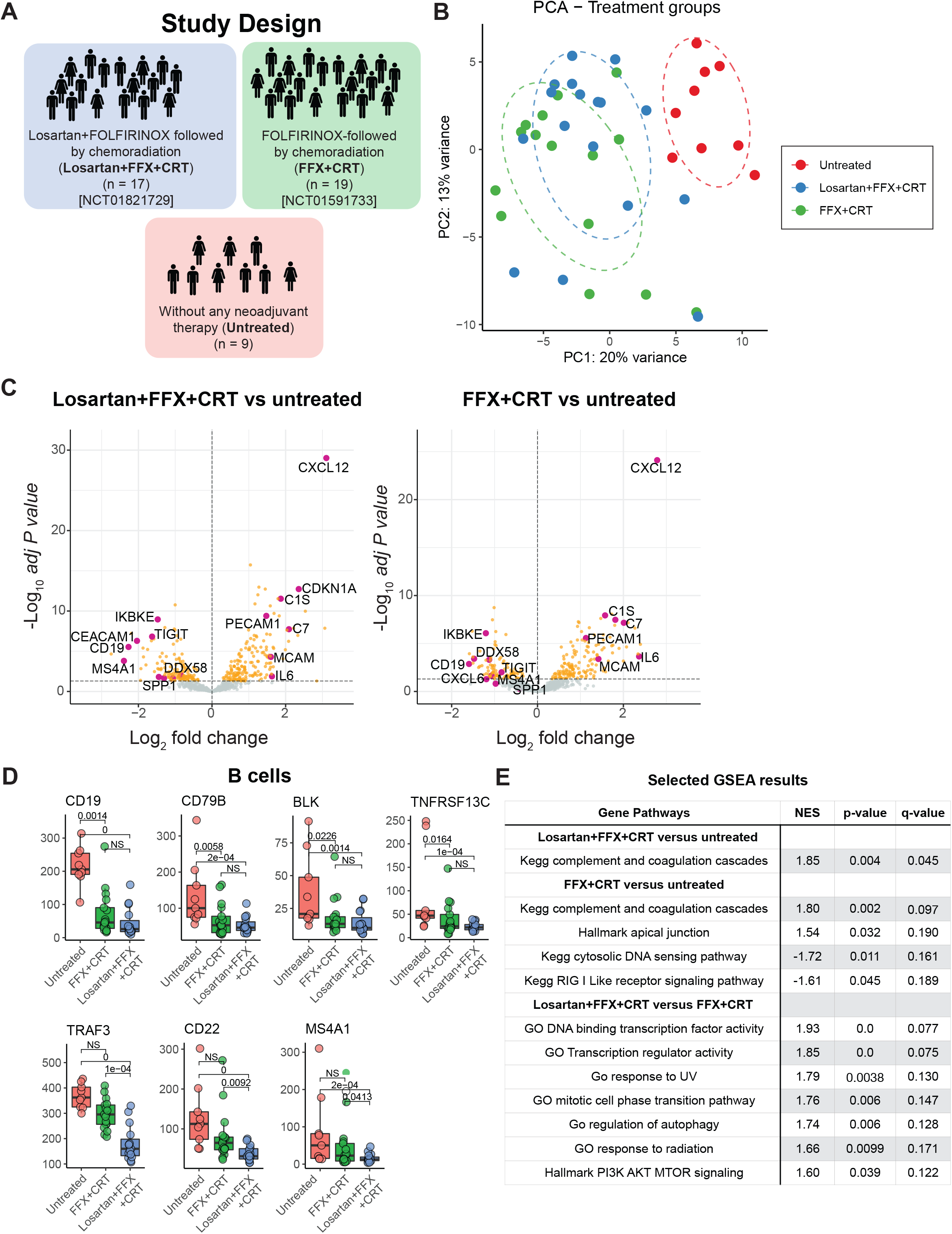
Differential gene expression between the 3 treatment groups (losartan+FFX+CRT n=15; FFX+CRT n=17; and untreated n=9). (**A**) Study design showing the sources of human tissues (**B**) PCA plots showing the clustering of all 3 groups. (**C**) Volcano plots of highly upregulated and downregulated genes between losartan+FFX+CRT vs untreated and losartan+FFX+CRT vs FFX+CRT. **(D)** Expression of B cell related genes in untreated, FFX+CRT and losartan+FFX+CRT samples. Adjusted p-values based on FDR set at 0.05. **(E)** Results of GSEA of pathways which are significantly different between losartan+FFX+CRT vs untreated, FFX+CRT vs untreated and losartan+FFX+CRT vs FFX+CRT.

### Analysis of gene expression in immune TME in resected samples

To characterize the effects and identify potential mechanisms of the activity of cytotoxic agents combined with losartan on the immune TME, we used the *nCounter PanCancer Immune Profiling Panel* of 730 genes (NanoString) to analyze the RNA extracted from paraffin embedded sections.

### Differential gene expression analysis and differential gene correlation analysis

The NanoString software package (nSolver) was used to import, process, export and analyze the nCounter raw data (RRID:SCR_003382). The *DESeq2* R package (RRID:SCR_000154) was used to determine differentially expressed genes (DEGs) between treatment groups. We used Benjamini & Hochberg method to control the False Discovery Rate (FDR) at 0.05. For principal component analysis (PCA), *prcomp* and *autoplot* functions were used from *stats* and *ggplot2* packages (RRID:SCR_014601), respectively. We made volcano plots and gene plots from the DEG list using the *EnhancedVolcano* and *ggplot2* packages (RRID:SCR_014601), respectively in R. For gene set enrichment analysis (GSEA), we used the GSEA application v4.2.3 [19]. We ran GSEA against the following MSigDB gene sets: GO, Hallmark, KEGG, C3 and C5 gene sets.

To correlate genes with one another and with respect to overall survival we used the Spearman Rank correlation (Prism Version 9 Software GraphPad). To determine whether gene signatures could stratify patient overall survival (< 36 months versus > 36 months) the expression of individual genes was transformed into Z-scores.

### Immunofluorescence

Immunofluorescence staining for CD4, FOXP3 and cytokeratin 19 was performed on surgically resected PDAC samples which were fixed in formalin and embedded in paraffin. Paraffin sections (5 µM thick) were baked for 3 h at 60 C then loaded into the BOND RX. Slides were deparaffinized in xylene and hydrated through graded alcohols. Antigen retrieval (Vector Citrate pH6 retrieval solution) was performed at pH 6.0 for 20 minutes at 98°C. Slides were incubated in CuSO_4_ for 90 min to block autofluorescence and blocked with 5% normal donkey serum. For the triple Foxp3, CD4, cytokeratin 19 stain, slides were incubated with FoxP3 (Biolegend 320102, 1:25) and CD4 (Abcam ab133616, 1:25) antibodies overnight at 4°C, followed by incubation with secondary antibodies (Jackson Immunoresearch) for 2 hours and cytokeratin 19-Alexa Fluor 488 antibody (Abcam ab192643, 1:50) overnight. All slides were counterstained with DAPI (NucBlue Fixed Cell ReadyProbes Reagent, Invitrogen), washed with deionized water, air dried, and mounted with ProLong Diamond Anti-fade Mountant (Invitrogen).

Imaging was performed with the Axio Scan.Z1 slide scanner (Zeiss) at 20x resolution. Images were analyzed using QuPath [20] and Python (RRID:SCR_001658). Cells were identified based on a positive DAPI signal, and each of the cell populations were classified as positive or negative based on a single intensity threshold on mean expression within the cell. Cells located in the tumor bed were included in the quantitative analysis. The mean number of positive or negative cells per mm^2^ of tissue was subsequently calculated and reported. The Wilcoxon test was conducted for each cell population on a per patient basis for each group. An alpha value of 0.05 was considered statistically significant. All analyses were performed using Prism Version 9 Software (GraphPad) and R Statistical Software (Foundation for Statistical Computing, Vienna, Austria).

## Results

Principal component analysis revealed that samples of patients not treated with neoadjuvant therapy clustered distinctly in comparison to losartan+FFX+CRT and/or FFX+CRT (**Fig. 1B**). Among the 730 genes tested, we detected 314 and 243 differentially expressed genes (DEGs) in losartan+FFX+CRT and FFX+CRT, respectively, versus control (**Supplementary Fig. 1**). Upon comparing the upregulated and downregulated genes in losartan+FFX+CRT and FFX+CRT with respect to untreated PDAC patients, we found that there were more genes overlapping between these two cohorts: 124 and 79 genes were commonly up- and downregulated, respectively, in losartan+FFX+CRT and FFX+CRT treated samples (**Supplementary Fig. 1**). The top 30 upregulated DEGs (fold-difference) in losartan+FFX+CRT and FFX+CRT versus untreated samples included cytokines / chemokines, complement factors, proliferation inhibition and blood vessel related genes (**Fig. 1C, Supplementary Table 1**). The top 30 downregulated DEGs in losartan+FFX+CRT versus untreated samples included immune checkpoints, pro-inflammatory genes and B cell related genes (**Fig. 1C, Supplementary Table 1**). The top 30 downregulated DEGs in FFX+CRT versus untreated samples included cytokines, RIG-I-like receptor signaling pathway and B cell related genes (**Fig. 1C, Supplementary Table 1**). FFX+CRT and losartan+FFX+CRT had similar as well as differential effects on the expression of B cell genes. FFX+CRT and losartan+FFX+CRT significantly downregulated the expression of B cell genes associated with BCR complex and signaling (*CD19, CD79B, BLK*) and survival (*TNFRSF13C*). In comparison to FFX+CRT, losartan+FFX+CRT induced a lower expression of B cell genes linked to B cell survival (*TRAF3, CD22*) and development and differentiation (*MSA41*) (**Fig. 1D**). The GSEA results revealed that FFX+CRT and losartan+FFX+CRT both enriched for increased complement and coagulation cascade gene sets. FFX+CRT also enriched for upregulation of the apical junction pathway and enriched for downregulation of cytosolic DNA sensing and RIG I like receptor pathways (**Fig. 1E**).

The direct comparison of losartan+FFX+CRT and FFX+CRT, revealed that 29 genes were downregulated, and 24 genes were upregulated (**Supplementary Fig. 2**). From GSEA, losartan+FFX+CRT versus FFX+CRT enriched for pathways involved in transcription factor activity, response to UV, response to radiation, cell cycle, regulation of autophagy and PI3K AKT MTOR signaling (**Fig 1E, Supplementary Table 2**). These findings reveal both similar and differential transcriptomic responses in PDAC after losartan+FFX+CRT versus FFX+CRT treatment.

### FFX+CRT with and without losartan increase the expression of genes involved in the development and differentiation of blood vessels, and the transendothelial migration of leukocytes

FFX+CRT and losartan+FFX+CRT increased the expression of genes linked to blood vessel maturation (*CDH5, THBS1, THBD, PDGFRB, ENG)* (**Fig. 2A**) and the transendothelial migration of leukocytes (*PECAM1, JAM3, MCAM, ICAM2, SELL, SELPLG*) (**Fig. 2B**). Losartan+FFX+CRT significantly reduced the expression of *TNFSF15*, while FFX did not affect the expression of *TNFSF15* (**Supplementary Fig. 3A**). The autocrine production of the TNFSF15 protein by endothelial cells inhibits endothelial cell proliferation, angiogenesis and tumor growth [21]. Taken together, these results suggest that FFX+CRT and losartan+FFX+CRT can stimulate angiogenesis and the normalization of the PDAC vasculature, as well as the transendothelial migration of leukocytes.

**Figure 2.**
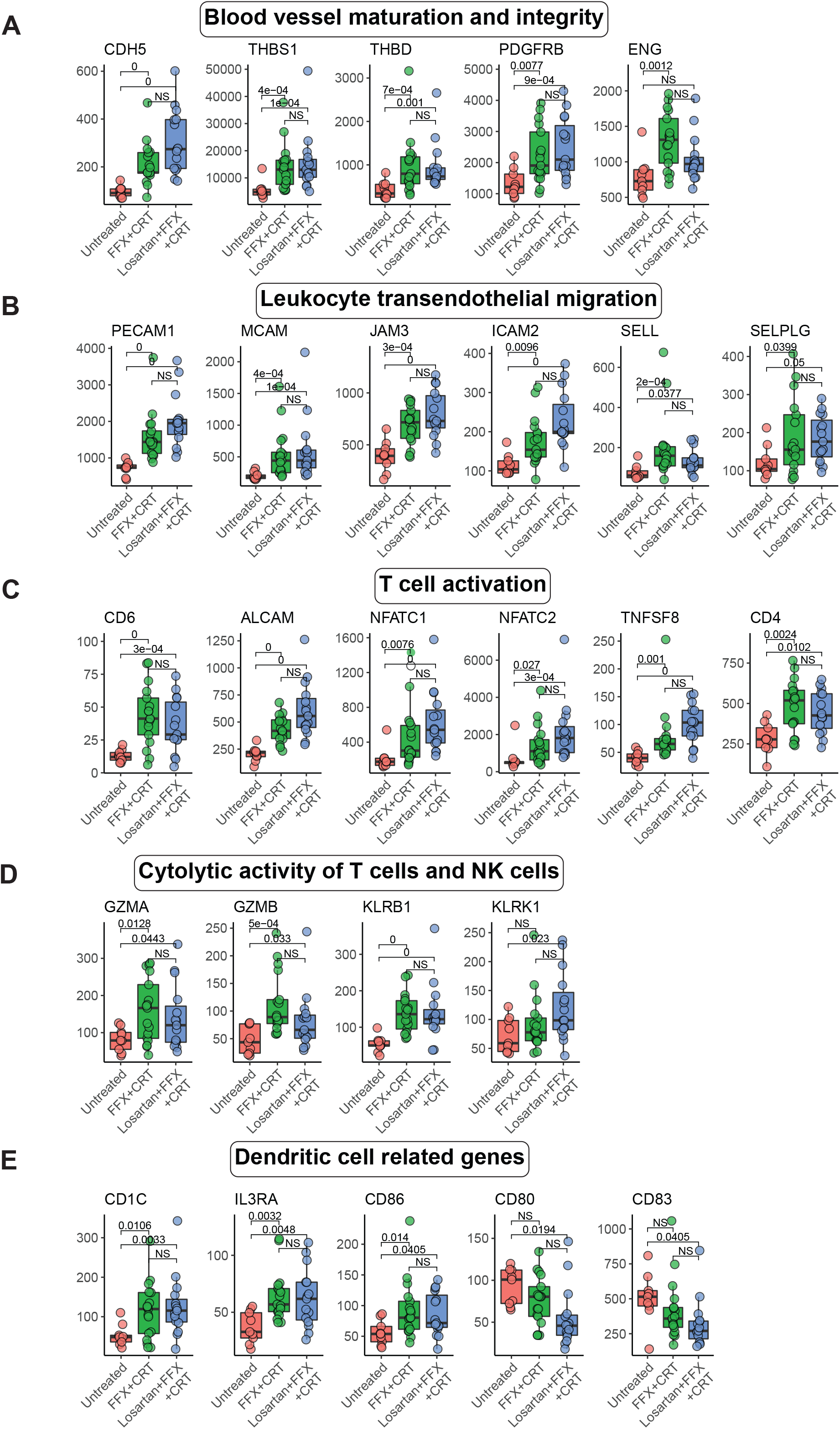
FFX+CRT and losartan+FFX+CRT enhance the expression of genes linked to the maturation of blood vessels, leukocyte transendothelial migration, T cell activation, cytolytic activity and dendritic cells. Effect of FFX+CRT and losartan+FFX+CRT on the expression of genes associated with blood vessel maturation and integrity (**A**), leukocyte transendothelial migration (**B**), T cell activation (**C**), cytolytic activity of T cells and NK cells (**D**) and dendritic cells (**E**). Losartan+FFX+CRT n=15; FFX+CRT n=17; and untreated n=9. Adjusted p-values based on FDR set at 0.05.

### FFX+CRT and losartan+FFX+CRT enhanced the expression of genes that play critical roles in T cell activation and immune cytolytic activity

FFX+CRT and losartan+FFX+CRT significantly reduced the expression of T helper (Th) Th1 (*IFNG, IL12A*) Th2 (*IL4, IL13*) and Th17 (*IL23A*) genes (**Supplementary Table 3**) but increased the expression of *CD6, ALCAM, TNFSF8, CD4, NFATC1 and NFATC2* (**Fig. 2C**). The cell adhesion molecule ALCAM binds to CD6 at the surface of T cells and promotes their activation and proliferation [22]. TNFSF8 / the CD30 ligand is expressed by macrophages, DCs and B cells and interacts with the CD30 receptor to promote the activation and proliferation of central memory T cells [23]. NFATC1 and NFATC2 are transcription factors involved in several biological processes including the formation of blood vessels, regulation of interaction of lymphocytes with endothelial cells and the activation of T cells. Losartan+FFX+CRT and FFX+CRT also increased the expression of cytolytic genes expressed by NK cells and T cells (*GZMA, GZMB, KLRB1)*, and losartan+FFX+CRT increased the expression of *KLRK1* (**Fig. 2D**).

### FFX+CRT and Losartan+FFX+CRT increase the expression of dendritic cell-selective genes

FFX+CRT and losartan+FFX+CRT significantly enhanced the expression of DC-associated genes [24], including *CD1C*, which is mostly found in MoDCs and conventional DC type 2 (cDC2s), and *IL3RA* (*CD123*), which is enriched in plasmacytoid DCs (pDCs) (**Fig. 2E**). Conversely, the expression of *CD1A*, typically over-expressed by cDC2s, was significantly down-regulated in both FFX+CRT and losartan+FFX+CRT-treated patients (**Supplementary Table 3**). *CD86*, a co-stimulatory factor that can be broadly expressed by various DC and macrophage types, was upregulated following either FFX+CRT or losartan+FFX+CRT treatment, whereas the expression of both *CD80* and *CD83* was lower in the losartan+FFX+CRT group (**Fig. 2E**). The expression of genes involved in the migration and maturation of DCs were also modulated by treatment. In comparison to untreated samples, both FFX+CRT and losartan+FFX+CRT increased the expression of *CCL17 and CSF1* (**Supplementary Fig. 3B**), which are respectively involved in the migration and maturation of DCs [25-27]. Interestingly, the expression of chemokine genes (*CCL3, CCL4*) which play roles in DC recruitment [28, 29] was higher in tumors treated with losartan+FFX+CRT (**Supplementary Fig. 3B**). The expression of *HMGB1* was also higher in samples treated with losartan+FFX+CRT (**Supplementary Fig. 3B**). The extracellular form of HMGB1 stimulates the maturation of DCs. These results suggest that FFX+CRT with or without losartan can stimulate the infiltration of specific DC subsets and maturation of DCs in PDAC.

### The expression of genes associated with the T cell receptor-CD3 complex and the migration of T cells and DCs to lymph nodes was higher in FFX+CRT than losartan+FFX+CRT treated samples

In comparison to losartan+FFX+CRT, in samples treated with FFX+CRT there was a higher expression of genes involved the phosphorylation/activation of the T cell receptor (TCR)-CD3 complex (*CD3G, LCK, ZAP70*) and in T cell development and activation (*SPN, IL7R*). In FFX+CRT treated samples there was also a higher expression of *CCR7*, which is not only involved in the migration of antigen-presenting DCs and antigen-specific and memory T cells to lymph nodes, but also defines an intratumoral DC state that provides survival and effector signals to tumor-specific T cells (**Fig. 3A**). Tumors from FFX+CRT-treated patients also showed a higher expression of *CCL21*, a chemokine gene that plays a role in the migration of *CCR7* expressing DCs to lymph nodes (**Supplementary Table 3**).

**Figure 3.**
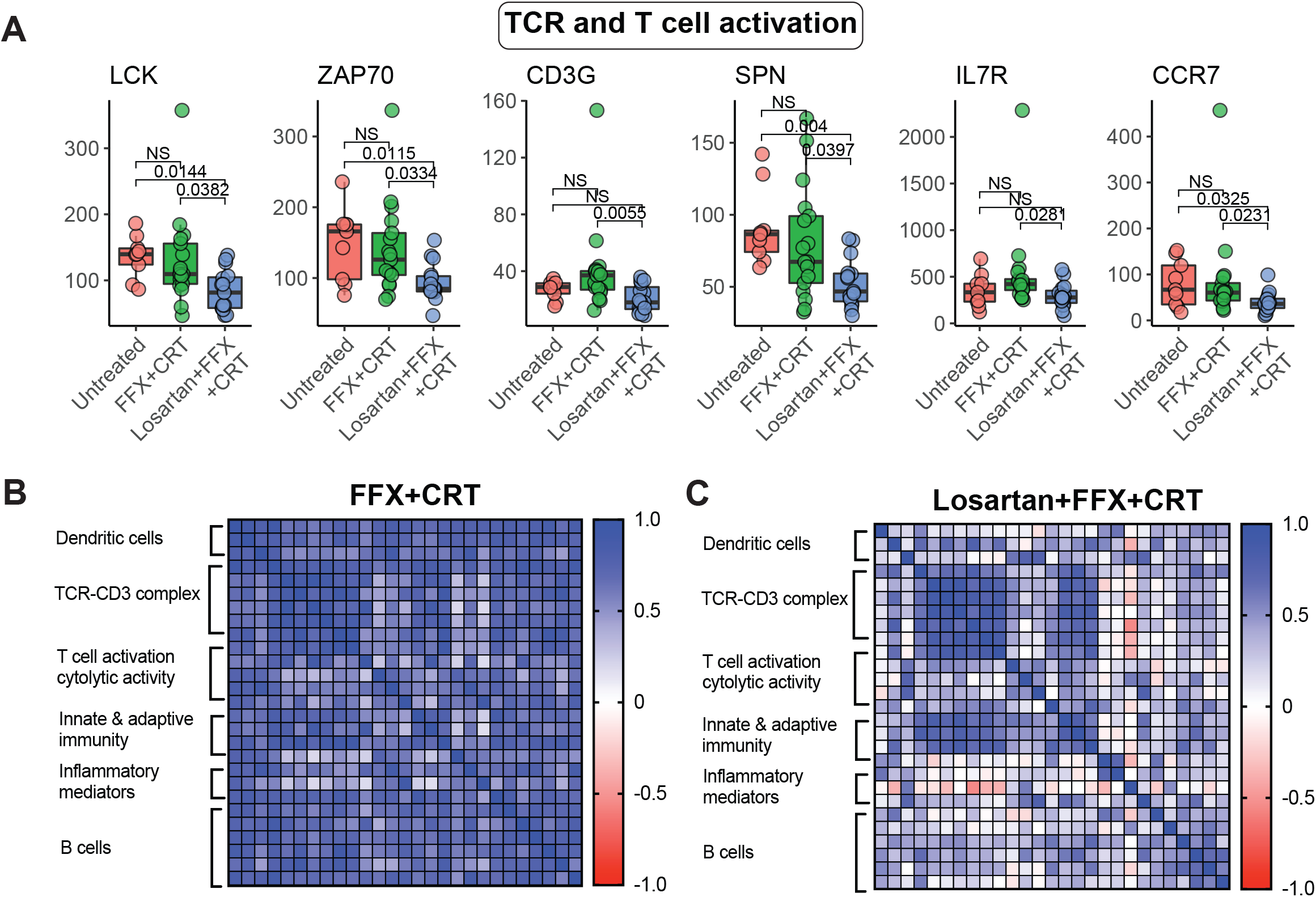
FFX+CRT vs losartan+FFX+CRT higher expression of genes that play a role in T cell receptor-CD3 complex and migration of T cells and DCs to lymph nodes. FFX+CRT increased expression of genes involved in TCR-CD3 complex, T cell activation and T cell and DC migration (**A**). Losartan+FFX+CRT n=15; FFX+CRT n=17; and untreated n=9. Adjusted p-values based on FDR set at 0.05. (**B**) Heatmap of Spearman rank correlation of genes associated with *CCR7* and *ZAP70* in tumor samples treated with FFX+CRT. (**C**) Heatmap of Spearman rank correlation of genes associated with *CCR7* and *ZAP70* in tumor samples treated with losartan+FFX+CRT.

In tumors treated with FFX+CRT we found a strong association between *CCR7* and *ZAP70* (Spearman R=0.917). CCR7 and ZAP70 were correlated with other DC-related genes, TCR-CD3 complex, T cell activation and cytolytic activity, innate and adaptive immunity, inflammatory mediators, and B cell related genes (**Fig. 3B, Supplementary Table 4**). In contrast, in tumors treated with losartan+FFX+CRT, except for the correlation of *CCR7* and *ZAP70* with genes of the TCR-CD3 complex, the correlation with other genes was weak (**Fig 3C**). These findings suggest that losartan+FFX+CRT reduces the activity of T cell receptor-CD3 complex and the migration of CCR7+ T cells or DCs.

### The addition of losartan to FFX+CRT reduces the expression of pro-invasion and upregulates genes and pathways involved in tumor suppression

Losartan+FFX+CRT versus FFX+CRT significantly decreased the expression of pro-invasion genes (*CEACAM6, CXCL16, ELK1*) (**Fig. 4A**) and increased the expression of tumor suppressor genes (*RORA, EP300*) (**Fig. 4B**). The retinoic acid receptor-related orphan receptor alpha (RORa) the protein product of *RORA* is involved in tumor suppression, and plays pro- and anti-inflammatory roles, which are context-dependent [30-32]. *RORA* in tumors treated with losartan+FFX+CRT correlated strongly with *NFATC4* (Spearman R=0.918) a gene also upregulated by losartan+FFX+CRT (**Supplementary Fig. 2**). The transcription factor Nuclear Factor of Activated T-Cells 4 is involved in the activation of T cells and endothelial cells [33, 34]. *RORA* and *NFATC4* correlated with tumor suppressor genes as well as blood vessels, DCs, MHCII, T cell activation and inflammation inhibition related genes, and were inversely associated with epithelial/cancer cell biomarkers (*EPCAM, CDH1*) (**Fig. 4C, Supplementary Table 5**). In contrast, in tumors treated with FFX+CRT there was no or weak correlations between the same gene sets (**Fig. 4D**). The expression of tumor suppressor genes and inverse correlation between cancer cell biomarkers and tumor suppressor genes, suggest that the addition of losartan to FFX+CRT reduces cancer cell proliferation / tumor progression. This is also supported by our GSEA results and immunofluorescence analysis of residual disease. The GSEA revealed an upregulation of the tumor suppressor p53 pathway (NES: 1.78; FDR q-value=0.044) (**Fig. 4E**) and circadian pathway (NES: 1.89; FDR q-value=0.090) (**Fig. 4F**). Circadian pathway genes regulate cell cycle check points as well as cell cycle progression [35]. Furthermore, we found significantly less residual disease (cytokeratin 19-positive cells) in lesions treated with losartan+FFX+CRT than FFX+CRT (**Fig. 4G**).

**Figure 4.**
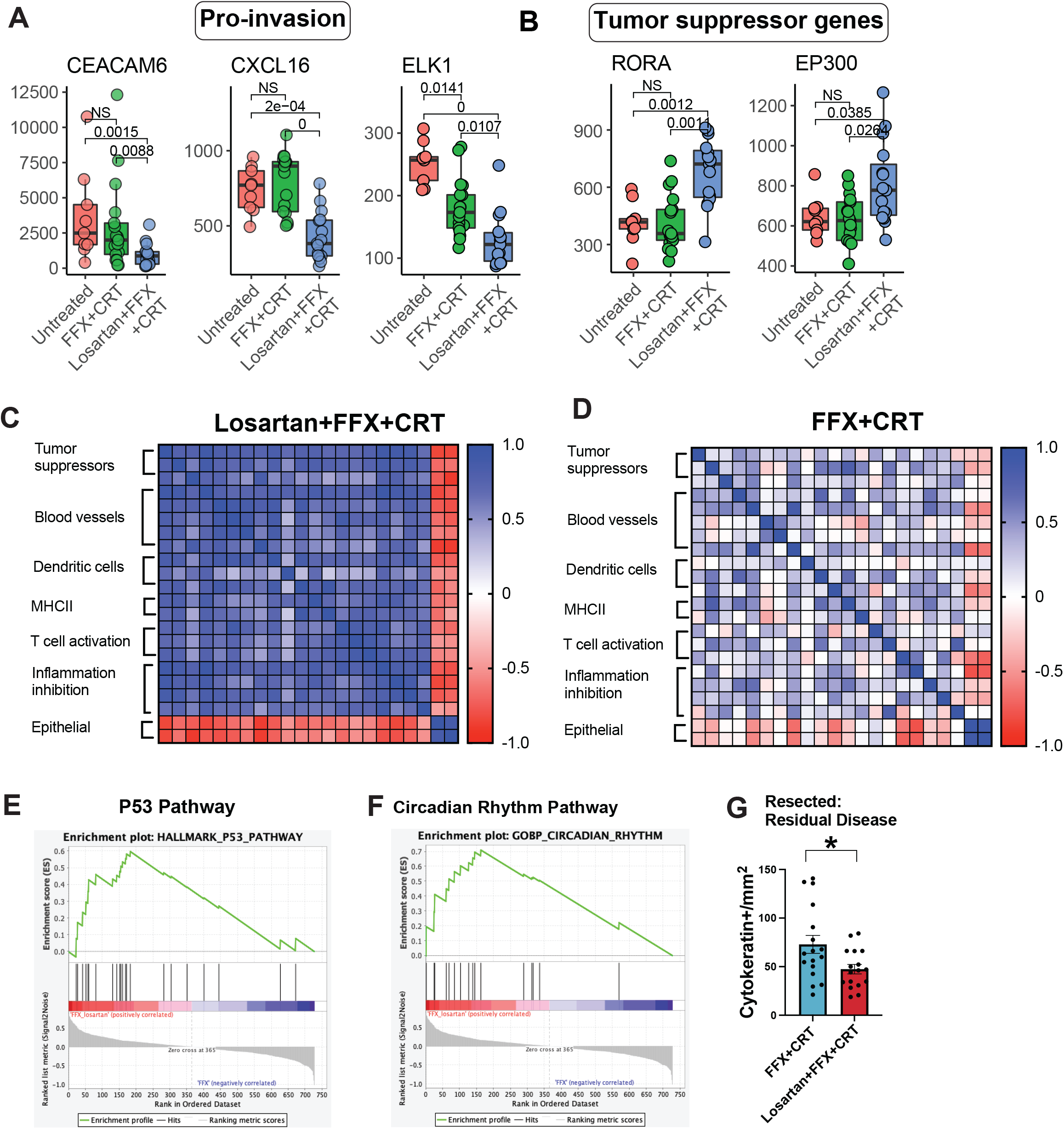
The addition of losartan to FFX+CRT decreased the expression of pro-invasion-associated genes and increrased the expression of tumor suppressor genes. **(A)** Pro-invasion genes. (**B**) Tumor suppressor genes. Losartan+FFX+CRT n=15; FFX+CRT n=17; and untreated n=9. Adjusted p-values based on FDR set at 0.05. (**C**) Heatmap of Spearman rank correlation of genes associated with *RORA* and *NFATC4* in tumor samples treated with losartan+FFX+CRT. (**D**) Heatmap of Spearman rank correlation of genes associated with *RORA* and *NFATC4* in tumor samples treated with FFX+CRT. (**E**) GSEA result of increased activity of P53 pathway in losartan+FFX+CRT versus FFX+CRT (NES: 1.78; FDR q-value=0.044). (**F**) GSEA result of increased activity of circadian rhythm pathway in losartan+FFX+CRT versus FFX+CRT (NES: 1.89; FDR q-value=0.090). (**G**) Quantitative analysis of residual disease (immunofluorescence of cytokeratin 19+ cells). * losartan+FFX+CRT N=17; FFX+CRT N=17; t-test p=0.025.

### The addition of losartan to FFX+CRT reduces the expression of immunosuppressive genes and Tregs in samples with less residual disease

The addition of losartan to FFX+CRT reduced the expression of M2 macrophage (*MARCO, APOE, CCR1*) related genes (**Fig. 5A**). The gene products of *APOE, MARCO and CCR1* are immunosuppressive in PDAC and other tumor types (see Discussion). FFX+CRT and losartan+FFX+CRT reduced the expression of the immune checkpoint gene *CEACAM1* (**Fig. 5B**). In comparison to untreated and FFX+CRT treated samples, losartan+FFX+CRT induced a significantly lower expression of the immune checkpoint *TIGIT* (**Fig. 5B**). The expression of the *FOXP3* gene—a transcription factor which regulates the activity of CD4^+^ Tregs which express TIGIT—was significantly lower in losartan+FFX+CRT-treated tumors than untreated tumors (**Fig. 5B**). We used immunofluorescence to determine the intratumoral infiltration of CD4^+^ T cells, and CD4^+^FOXP3^+^ Tregs (**Fig 5C**). In PDAC lesions treated with FFX+CRT or losartan+FFX+CRT, there was no difference in number of CD4^+^ and CD4^+^FOXP3^+^ cells (Tregs) between PDAC lesions treated with losartan+FFX+CRT versus FFX+CRT (**Fig. 5D, E**). However, in resected samples from patients treated with losartan+FFX+CRT we found significantly fewer Tregs in PDAC lesions with a complete/near complete (0/1) versus partial/poor or no response (2/3) (as grouped by the College of American Pathologists tumor regression grading system in PDAC after neoadjuvant therapy [36]) (**Fig. 5F**). In resected samples from patients treated with FFX+CRT there was no difference in the number of Tregs between PDAC lesions with a complete/near complete (0/1) versus partial/poor or no response (2/3) (**Supplementary Fig. 4**). These results suggest that the addition of losartan to FFX+CRT can decrease Tregs in patients with a better pathological response.

**Figure 5.**
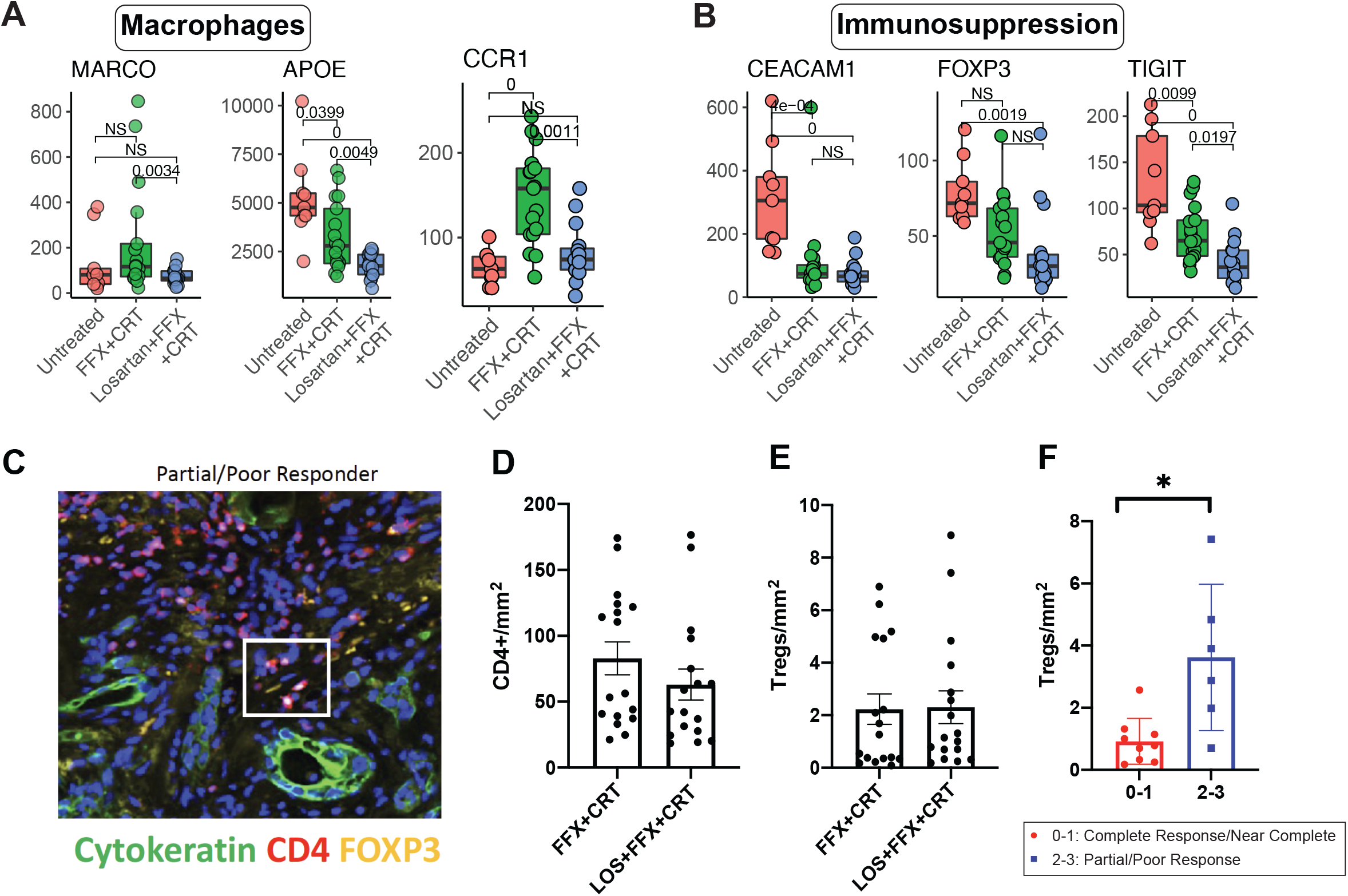
Losartan+FFX+CRT reduces the expression of immunosuppressive genes and regulatory T cells in samples with less residual disease. Losartan+FFX+CRT vs FFX+CRT induce a greater down-regulation of macrophage related genes (**A**). Losartan+FFX+CRT and FFX+CRT induced a significant down-regulation of the immune checkpoint *CEACAM 1*, while the addition of losartan to FFX+CRT induce a greater down-regulation of *TIGIT* and *FOXP3*. Losartan+FFX+CRT n=15; FFX+CRT n=17; and untreated n=9. Adjusted p-values based on FDR set at 0.05. (**B**). Immunofluorescence staining of CD4+ T cells, CD4+FOXP3+ Tregs and cytokeratin-19 positive cells (**C**). Quantitative analysis of CD4+ T cells (**D**), and CD4+FOXP3+ Tregs in PDAC lesions treated with losartan+FFX+CRT and FFX+CRT (**E**). Quantitative analysis of CD4+FOXP3+ Tregs in PDAC lesions treated with losartan+FFX+CRT comparing the complete/near complete versus poor/no response. * Losartan+FFX+CRT N=15, t-test p<0.01 (**F**).

### In patients treated with losartan+FFX+CRT the lower expression of immunosuppression and invasion / proliferation genes is associated with improved overall survival

We assessed whether gene sets could identify differences in the immune and non-immune TME between low (< 36 months) versus high (> 36 months) OS. To perform this analysis, we included genes positively (Spearman R ≥ 0.5) or negatively correlated (Spearman R ≥ -0.5) with OS (**Supplementary Table 6 and 7**). For samples treated with losartan+FFX+CRT, a gene set which included immunosuppression and invasion / proliferation genes stratified OS (**Fig 6A, Supplementary Table 8**). Gene sets which included only immunosuppression and invasion genes (**Fig 6B**), or only immunosuppression or invasion / proliferation genes could also stratify OS (**Fig. 6C, Supplementary Table 8**). The gene set positively associated with OS – which included 30 genes – did not stratify with OS, but smaller grouping of genes which included blood vessel maturation, transendothelial migration, DCs and tumor suppression related genes stratified OS (**Fig 6D, Supplementary Table 8)**. For samples treated with FFX+CRT, the gene sets negatively or positively associated with OS stratified OS (**Fig 6E – 6H**). DC and antigen presentation related genes provided a better stratification of OS (**Fig. 6H, Supplementary Table 8)** than the 17 gene set (**Fig. 6H**) which also included B cell related genes. Thus, for patients treated with losartan+FFX+CRT the lower expression of immunosuppression and invasion related genes, and higher expression of blood vessel and DC related genes is associated with improved OS.

**Figure 6.**
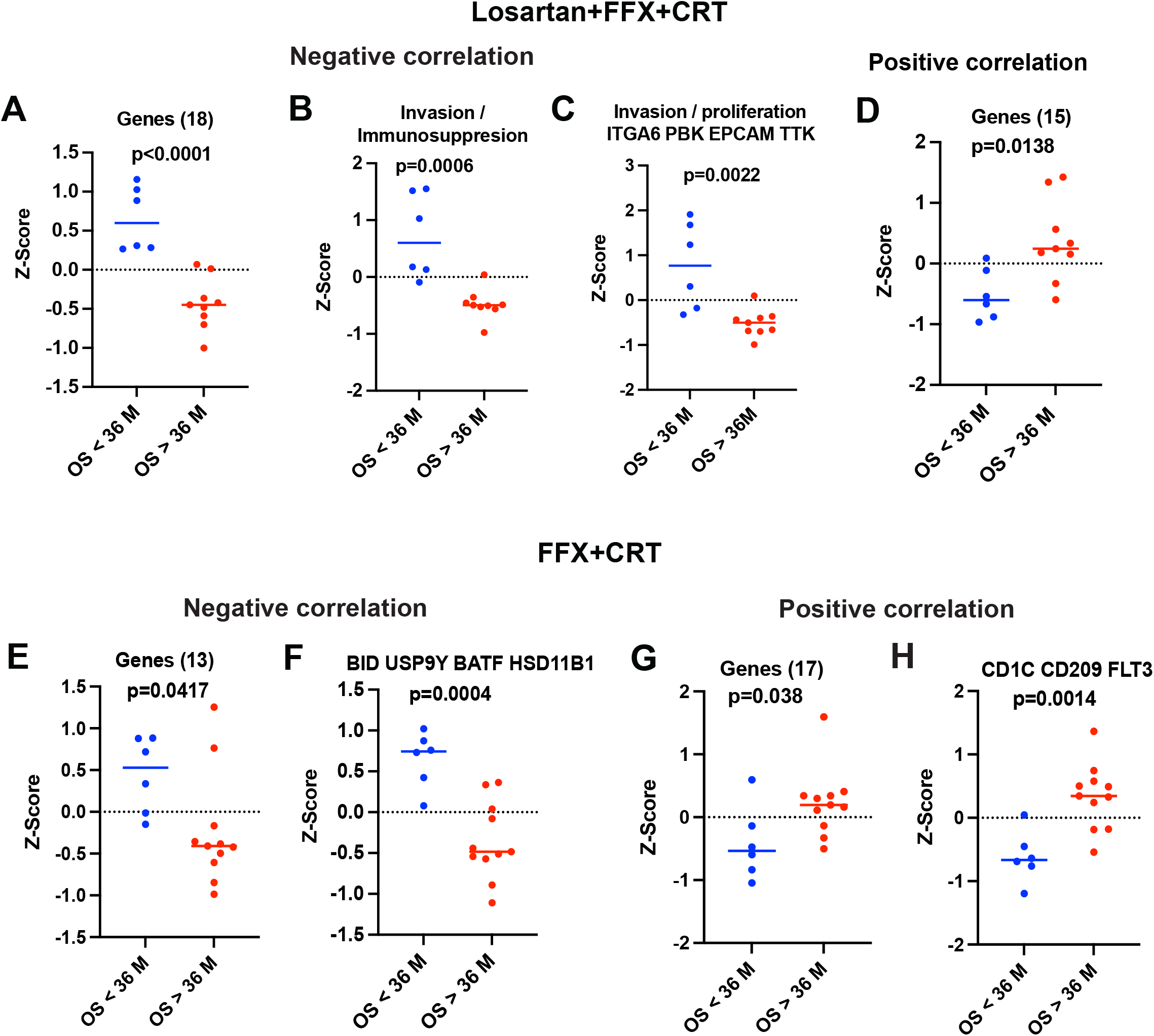
Gene set stratification of overall survival (OS) for patients treated with FFX+losartan+CRT and FFX+CRT. For patients treated with FFX+losartan+CRT genes sets which included immunosuppression and invasion / proliferation (**A**), or only invasion / immunosuppression (**B**) and only invasion / proliferation genes (**C**) DCs, blood vessel maturation, transendothelial migration and tumor suppression related genes stratified OS (**D**). For patients treated with FFX+CRT gene sets negatively (**E, F)** or positively (**G, H)** associated with OS stratified OS. Losartan+FFX+CRT n=15; FFX+CRT n=17. P values based on t-test.

We also assessed whether a publicly available DC gene signature is associated with OS [37]. Our analysis revealed that the DC gene signature stratified the OS of patients treated with FFX+CRT, especially when the *HSD11B1* gene was not included (**Supplementary Table 9**). The expression of *HSD11B1* in samples treated with FFX+CRT is inversely correlated with OS (**Supplementary Table 7**). The elevated expression of 11b-hydroxysteroid dehydrogenase type 1 glucocorticoid – the gene product of *HSD11B1* – in DCs inhibits their activity [38]. *HSD11B1* overexpression has been associated with poor prognosis in some cancer types [39]. In contrast, in patients treated with losartan+FFX+CRT the same gene signature [37] did not associate with OS (**Supplementary Table 9**).

## Discussion

Our results show that FFX+CRT and losartan+FFX+CRT induced several changes in borderline PDAC and LAPC samples, respectively. These included increases in gene expression levels for factors involved in 1) the maturation of blood vessels and transendothelial migration of leukocytes, 2) activation of T cells, 3) cytolytic activity of T cells and NK cells, and 4) DC activity. The addition of losartan to FFX treatment further 1) reduced the expression of genes associated with immunosuppression and invasion and 2) reduced Tregs in resected samples with a complete/near complete pathological response.

A detailed analysis showed that addition of losartan to FFX+CRT reduced the expression of pro-invasion genes (*CEACAM6, CXCL16, ELK1*), as well as M2 macrophage-associated genes (*APOE, MARCO, CCR1*) involved in tumor progression and immunosuppression. In pancreatic cancer models CEACAM6, CXCL16, ELK1 can stimulate tumor cell proliferation and invasion [40-43]. In PDAC, APOE – which is expressed by fibroblasts, macrophages, and plasma cells – stimulates the infiltration of MDSCs which suppresses the infiltration of CD4+ and CD8+ T cells [3]. The expression of CCR1 by MDSCs and macrophages in PDAC has also been linked to immunosuppression [44]. In a PDAC model, MARCO promoted tumor progression and metastasis [45], while another report showed that high expression of *MARCO* is an independent indicator of poor prognosis in PDAC [46]. Targeting of MARCO in lung cancer models reduced the activity of Tregs and potentiated the cytolytic and anti-tumor activity of T cells and NK cells [47]. Our results also revealed a lower expression of pro-invasion and immunosuppression-related genes in resected samples of PDAC patients with superior OS (**Fig 6A-C, Supplementary Table 8**). We have shown in a transcriptome analysis that the chronic administration of the ASI lisinopril in patients with PDAC reduces pro-invasive pathways (ECM / receptor interaction, WNT and Notch signaling) and expression of pro-invasion genes [11]. Thus, the decrease expression of pro-invasion and M2 macrophages genes suggest that the addition of losartan to FFX+CRT produces a PDAC TME that attenuates invasion and immunosuppression.

FFX+CRT reduced the expression of the immune checkpoints *TIGIT* and *CEACAM1*, while the addition of losartan to FFX+CRT induced a significantly greater reduction in *TIGIT*, reduced the expression of *FOXP3*, and in resected PDAC lesions with less residual disease there was less Tregs. TIGIT is expressed by a subset of

Tregs that suppress the activity of Th1 and Th17 cells [48, 49], while in human PDAC TIGIT is expressed by Tregs and CD8+ T cells [7]. Inhibition of TIGIT combined with PD-1 blockade and CD40 activation potently induced tumor regression in orthotopic PDAC models [50]. Interestingly, TIGIT inhibition increased the production of IFND and proliferation of CD8^+^ T cells isolated from the blood of PDAC patients treated with FFX but did not affect the activity of CD8^+^ T cells isolated before the administration of FFX [51]. These findings suggest that the losartan-induced reduction of Tregs and down-regulation of TIGIT could potentiate the anti-tumor activity of FFX+CRT in PDAC.

Losartan+FFX+CRT significantly reduced the expression of several B-cell related genes. The AT1 receptor is expressed by 69% of B cells [52] and in a model of inflammation losartan significantly reduced the viability of B cells [53]. Furthermore, stereotactic body radiation therapy depletes circulating CD19^+^ B cells to a greater extent than T cell subtypes [54]. These findings suggest that losartan combined with radiation could significantly impair the development and viability of B cells in PDAC.

In PDAC the paucity of conventional DCs (cDC1 and cDC2) [5, 6, 55] in the TME impairs immune surveillance and accelerates tumor progression [5, 6]. The production of granulocyte colony stimulating factor and IL-6 in mouse models of PDAC and human PDAC impairs the development of cDC1s [6, 56]. The Fms-related Receptor Tyrosine Kinase 3 Ligand (Flt3L) combined with CD40 agonist antibody significantly increased the infiltration of cDC1s and cDC2s in PDAC, but while the combination inhibited tumor growth it did not induce tumor regressions [5]. In contrast, the addition of radiation to Flt3L and CD40 activation induced the regression of PDAC tumors. Here we show that FFX+CRT with and without losartan increased the expression of DC-related genes *CD1C* and *IL3RA*, and in FFX+CRT-treated samples the expression of *CD1C* and *CD209 –* which can be expressed by DCs or macrophages – was associated with improved OS. Gene sets that included *IL3RA* and *CSF1* were also associated with improved OS in samples treated with losartan+FFX+CRT. In another study which analyzed resected samples from untreated PDAC patients, the higher gene expression of *CD209* and *CD1C* was also correlated with improved survival [57]. While we found high levels of *CD1C* (cDC2) and *IL3RA* (pDCs) in PDAC patients with improved survival, it will be significant in mechanistic studies to assess the specific role of cDC2 and pDCs in PDAC immunosurveillance. Of significance, cDC2 vaccination in mice reduced intratumoral MDSCs, reprogrammed M2 macrophages and inhibited tumor growth in a colon carcinoma model [58]. Furthermore, in another study Treg depletion reduced the suppression of cDC2 and potentiated the anti-tumor activity of conventional CD4^+^ T cells [59].

Neoadjuvant FFX can increase the infiltration of effector T cells and decrease immunosuppressive cells in patients with PDAC [17]. The results of our transcriptome analysis showing an increase in expression of genes linked to blood vessel maturation and transendothelial migration of leukocytes is consistent with this improvement in T cell infiltration induced by FFX or FFX combined with CRT. The maturation of blood vessels has been associated with the increased infiltration of CD4+ and CD8+ T cells and efficacy of immunotherapy [60-62]. PECAM-1, MCAM, vascular I-CAM2 and VE-cadherin the gene product of *CDH5* play significant roles in the transendothelial migration of leukocytes [63]. For example, the targeting of endothelial VE-cadherin enhanced the intratumoral infiltration of T cells and T cell-mediated immunotherapy [64].

Our study has several limitations. Resected surgical tumors may not be the optimal control baseline for borderline or locally advanced disease. However, pre-clinical studies in mice as well as studies which compared benign lesions versus invasive PDAC have shown that the TME of PDAC is immunosuppressive even at early stages of the disease [6, 55]. Another limitation is that samples belonging to the losartan+FFX+CRT and FFX+CRT groups were not obtained from a randomized clinical trial. Hence, there might be confounding factors which could influence the transcriptomic differences observed here. Finally, the NanoString approach is limited to a set of genes included in the panel. These limitations notwithstanding, our transcriptomic analysis performed with RNA extracted from FFPE sections and the NanoString platform provides novel insights on the effects of FFX+CRT versus losartan+FFX+CRT on the expression and functional activity of invasion- and immune-related genes in the PDAC TME.

In conclusion, our findings show that addition of losartan to FFX+CRT reduced pro-invasion and immunosuppression in PDAC, which was associated with improved treatment outcomes in patients with LAPC. The decreased expression of pro-invasion and M2 macrophages genes, combined with the decreased expression of *TIGIT* and the number of Tregs in lesions with less residual disease suggest that the addition of losartan to FFX+CRT further reprograms the PDACs toward reduced invasion and TME immunosuppression. Our findings offer a potential explanation for the OS benefit observed in a retrospective analysis of GI patients who were receiving ASIs along with immune-checkpoint blockers [65].

## Data Availability

All data produced in the present study are available upon reasonable request to the authors

## Acknowledgements

We thank Carolyn Smith for technical assistance. We would also like to acknowledge the contribution of Drs. Ivy X. Chen and Mei R. Ng with immunohistochemistry protocols.

## Supplementary Tables

**Supplementary Table 1**. Top 30 upregulated and 30 downregulated genes losartan+FFX+CRT vs untreated and FFX+CRT vs untreated.

**Supplementary Table 2**. Gene Set Enrichment Analysis results of losartan+FFX+CRT vs FFX+CRT.

**Supplementary Table 3**. DEGs losartan+FFX+CRT vs untreated and FFX+CRT vs untreated.

**Supplementary Table 4**. Genes included in panels B and C of Figure 3.

**Supplementary Table 5**. Genes included in panels C and D of Figure 4.

**Supplementary Table 6**. Losartan+FFX+CRT correlation of individual genes with overall survival.

**Supplementary Table 7**. FFX+CRT correlation of individual genes with overall survival.

**Supplementary Table 8**. Gene grouping association with overall survival.

**Supplementary Table 9**. Gene signature association with overall survival.

## Supplementary Figure Legends

**Supplementary Figure 1. Venn plots showing the overlap of upregulated and downregulated genes between losartan+FFX+CRT vs untreated tumors and FFX+CRT vs untreated tumors**.

**Supplementary figure 2. Heatmap showing 54 DEGs and their expression in each patient**. We grouped rows based on treatment category and columns based on hierarchical clustering.

**Supplementary figure 3. Effect of losartan+FFX+CRT and FFX+CRT on genes involved in angiogenesis and the migration and maturation of DCs**. Losartan+FFX+CRT induced a greater down-regulation of

*TNFSF15* (**A**). In comparison to untreated samples, both FFX+CRT and losartan+FFX+CRT increased the expression of *CCL17 and CSF1* (**B**). The expression of chemokine genes (*CCL3, CCL4*) was higher in samples treated with losartan+FFX+CRT than untreated samples (**B**). In comparison to FFX+CRT, losartan+FFX+CRT significantly increased the expression of *HMGB1* (**B**). Losartan+FFX+CRT n=15; FFX+CRT n=17; and untreated n=9. Adjusted p-values based on FDR set at 0.05.

**Supplementary figure 4. Effect of FFX+CRT on CD4+FOXP3+ Tregs in PDAC lesions with a complete/near complete versus poor/no response**. FFX+CRT N=17.

## References

1. Sideras, K., et al., Role of the immune system in pancreatic cancer progression and immune modulating treatment strategies. Cancer Treat Rev, 2014. 40(4): p. 513–22.

2. Bazhin, A.V., et al., Two immune faces of pancreatic adenocarcinoma: possible implication for immunotherapy. Cancer Immunol Immunother, 2014. 63(1): p. 59–65.

3. Kemp, S.B., et al., Apolipoprotein E Promotes Immune Suppression in Pancreatic Cancer through NF-kappaB-Mediated Production of CXCL1. Cancer Res, 2021. 81(16): p. 4305–4318.

4. Huber, M., et al., The Immune Microenvironment in Pancreatic Cancer. Int J Mol Sci, 2020. 21(19).

5. Hegde, S., et al., Dendritic Cell Paucity Leads to Dysfunctional Immune Surveillance in Pancreatic Cancer. Cancer Cell, 2020. 37(3): p. 289–307 e9.

6. Lin, J.H., et al., Type 1 conventional dendritic cells are systemically dysregulated early in pancreatic carcinogenesis. J Exp Med, 2020. 217(8).

7. Sivakumar, S., et al., Activated Regulatory T-Cells, Dysfunctional and Senescent T-Cells Hinder the Immunity in Pancreatic Cancer. Cancers (Basel), 2021. 13(8).

8. Chauhan, V.P., et al., Angiotensin inhibition enhances drug delivery and potentiates chemotherapy by decompressing tumour blood vessels. Nat Commun, 2013. 4: p. 2516.

9. Stylianopoulos, T., et al., Causes, consequences, and remedies for growth-induced solid stress in murine and human tumors. Proc Natl Acad Sci U S A, 2012. 109(38): p. 15101–8.

10. Murphy, J.E., et al., Total Neoadjuvant Therapy With FOLFIRINOX in Combination With Losartan Followed by Chemoradiotherapy for Locally Advanced Pancreatic Cancer: A Phase 2 Clinical Trial. JAMA Oncol, 2019. 5(7): p. 1020–1027.

11. Liu, H., et al., Use of Angiotensin System Inhibitors Is Associated with Immune Activation and Longer Survival in Nonmetastatic Pancreatic Ductal Adenocarcinoma. Clin Cancer Res, 2017. 23(19): p. 5959–5969.

12. Labiano, S., A. Palazon, and I. Melero, Immune response regulation in the tumor microenvironment by hypoxia. Semin Oncol, 2015. 42(3): p. 378–86.

13. Cortez-Retamozo, V., et al., Angiotensin II drives the production of tumor-promoting macrophages. Immunity, 2013. 38(2): p. 296–308.

14. Ruffell, B., et al., Macrophage IL-10 blocks CD8+ T cell-dependent responses to chemotherapy by suppressing IL-12 expression in intratumoral dendritic cells. Cancer Cell, 2014. 26(5): p. 623–37.

15. Pfirschke, C., et al., Immunogenic Chemotherapy Sensitizes Tumors to Checkpoint Blockade Therapy. Immunity, 2016. 44(2): p. 343–54.

16. Engblom, C., C. Pfirschke, and M.J. Pittet, The role of myeloid cells in cancer therapies. Nat Rev Cancer, 2016. 16(7): p. 447–62.

17. Michelakos, T., et al., Tumor microenvironment immune response in pancreatic ductal adenocarcinoma patients treated with neoadjuvant therapy. J Natl Cancer Inst, 2021. 113: p. 182-191.

18. Peng, H., et al., Neoadjuvant FOLFIRINOX Therapy Is Associated with Increased Effector T Cells and Reduced Suppressor Cells in Patients with Pancreatic Cancer. Clin Cancer Res, 2021. 27(24): p. 6761–6771.

19. Subramanian, A., et al., Gene set enrichment analysis: a knowledge-based approach for interpreting genome-wide expression profiles. Proc Natl Acad Sci U S A, 2005. 102(43): p. 15545–50.

20. Bankhead, P., et al., QuPath: Open source software for digital pathology image analysis. Sci Rep, 2017. 7(1): p. 16878.

21. Zhou, J., et al., LITAF and TNFSF15, two downstream targets of AMPK, exert inhibitory effects on tumor growth. Oncogene, 2011. 30(16): p. 1892–900.

22. Zimmerman, A.W., et al., Long-term engagement of CD6 and ALCAM is essential for T-cell proliferation induced by dendritic cells. Blood, 2006. 107(8): p. 3212–20.

23. Nishimura, H., et al., A novel role of CD30/CD30 ligand signaling in the generation of long-lived memory CD8+ T cells. J Immunol, 2005. 175(7): p. 4627–34.

24. Gerhard, G.M., et al., Tumor-infiltrating dendritic cell states are conserved across solid human cancers. J Exp Med, 2021. 218(1).

25. MacDonald, K.P., et al., The colony-stimulating factor 1 receptor is expressed on dendritic cells during differentiation and regulates their expansion. J Immunol, 2005. 175(3): p. 1399–405.

26. Wiechers, C., et al., Lymph node stromal cells support the maturation of pre-DCs into cDC-like cells via colony-stimulating factor 1. Immunology, 2022.

27. Stutte, S., et al., Requirement of CCL17 for CCR7-and CXCR4-dependent migration of cutaneous dendritic cells. Proc Natl Acad Sci U S A, 2010. 107(19): p. 8736–41.

28. Williford, J.M., et al., Recruitment of CD103(+) dendritic cells via tumor-targeted chemokine delivery enhances efficacy of checkpoint inhibitor immunotherapy. Sci Adv, 2019. 5(12): p. eaay1357.

29. Allen, F., et al., CCL3 Enhances Antitumor Immune Priming in the Lymph Node via IFNgamma with Dependency on Natural Killer Cells. Front Immunol, 2017. 8: p. 1390.

30. Hams, E., et al., Functions for Retinoic Acid-Related Orphan Receptor Alpha (RORalpha) in the Activation of Macrophages During Lipopolysaccharide-Induced Septic Shock. Front Immunol, 2021. 12: p. 647329.

31. Ma, X., et al., CircGSK3B promotes RORA expression and suppresses gastric cancer progression through the prevention of EZH2 trans-inhibition. J Exp Clin Cancer Res, 2021. 40(1): p. 330.

32. Lee, J.M., H. Kim, and S.H. Baek, Unraveling the physiological roles of retinoic acid receptor-related orphan receptor alpha. Exp Mol Med, 2021. 53(9): p. 1278–1286.

33. Muller, M.R. and A. Rao, NFAT, immunity and cancer: a transcription factor comes of age. Nat Rev Immunol, 2010. 10(9): p. 645–56.

34. Yu, B.X., et al., Inhibition of Orai1-mediated Ca(2+) entry limits endothelial cell inflammation by suppressing calcineurin-NFATc4 signaling pathway. Biochem Biophys Res Commun, 2018. 495(2): p. 1864–1870.

35. Farshadi, E., G.T.J. van der Horst, and I. Chaves, Molecular Links between the Circadian Clock and the Cell Cycle. J Mol Biol, 2020. 432(12): p. 3515–3524.

36. Cacciato Insilla, A., et al., Tumor Regression Grading Assessment in Locally Advanced Pancreatic Cancer After Neoadjuvant FOLFIRINOX: Interobserver Agreement and Prognostic Implications. Front Oncol, 2020. 10: p. 64.

37. Bindea, G., et al., Spatiotemporal dynamics of intratumoral immune cells reveal the immune landscape in human cancer. Immunity, 2013. 39(4): p. 782–95.

38. Soulier, A., et al., Cell-intrinsic regulation of murine dendritic cell function and survival by prereceptor amplification of glucocorticoid. Blood, 2013. 122(19): p. 3288–97.

39. Li, C.F., et al., Hydroxysteroid 11-Beta Dehydrogenase 1 Overexpression with Copy-Number Gain and Missense Mutations in Primary Gastrointestinal Stromal Tumors. J Clin Med, 2018. 7(11).

40. Chalabi-Dchar, M., et al., Loss of Somatostatin Receptor Subtype 2 Promotes Growth of KRAS-Induced Pancreatic Tumors in Mice by Activating PI3K Signaling and Overexpression of CXCL16. Gastroenterology, 2015. 148(7): p. 1452–65.

41. Yan, Q., et al., ELK1 Enhances Pancreatic Cancer Progression Via LGMN and Correlates with Poor Prognosis. Front Mol Biosci, 2021. 8: p. 764900.

42. Chen, J., et al., CEACAM6 induces epithelial-mesenchymal transition and mediates invasion and metastasis in pancreatic cancer. Int J Oncol, 2013. 43(3): p. 877–85.

43. Cheng, T.M., et al., Single domain antibody against carcinoembryonic antigen-related cell adhesion molecule 6 (CEACAM6) inhibits proliferation, migration, invasion and angiogenesis of pancreatic cancer cells. Eur J Cancer, 2014. 50(4): p. 713–21.

44. Zhang, Y., et al., Regulatory T-cell Depletion Alters the Tumor Microenvironment and Accelerates Pancreatic Carcinogenesis. Cancer Discov, 2020. 10(3): p. 422–439.

45. Neyen, C., et al., Macrophage scavenger receptor a promotes tumor progression in murine models of ovarian and pancreatic cancer. J Immunol, 2013. 190(7): p. 3798–805.

46. Shi, B., et al., The Scavenger Receptor MARCO Expressed by Tumor-Associated Macrophages Are Highly Associated With Poor Pancreatic Cancer Prognosis. Front Oncol, 2021. 11: p. 771488.

47. La Fleur, L., et al., Targeting MARCO and IL37R on Immunosuppressive Macrophages in Lung Cancer Blocks Regulatory T Cells and Supports Cytotoxic Lymphocyte Function. Cancer Res, 2021. 81(4): p. 956–967.

48. Joller, N., et al., Treg cells expressing the coinhibitory molecule TIGIT selectively inhibit proinflammatory Th1 and Th17 cell responses. Immunity, 2014. 40(4): p. 569–81.

49. Kurtulus, S., et al., TIGIT predominantly regulates the immune response via regulatory T cells. J Clin Invest, 2015. 125(11): p. 4053–62.

50. Freed-Pastor, W.A., et al., The CD155/TIGIT axis promotes and maintains immune evasion in neoantigen-expressing pancreatic cancer. Cancer Cell, 2021. 39(10): p. 1342–1360 e14.

51. Jin, H.S., et al., CD226(hi)CD8(+) T Cells Are a Prerequisite for Anti-TIGIT Immunotherapy. Cancer Immunol Res, 2020. 8(7): p. 912–925.

52. Rasini, E., et al., Angiotensin II type 1 receptor expression on human leukocyte subsets: a flow cytometric and RT-PCR study. Regul Pept, 2006. 134(2-3): p. 69–74.

53. Wang, X., et al., Losartan suppresses the inflammatory response in collagen-induced arthritis by inhibiting the MAPK and NF-kappaB pathways in B and T cells. Inflammopharmacology, 2019. 27(3): p. 487–502.

54. Zhuang, Y., et al., Association Between Circulating Lymphocyte Populations and Outcome After Stereotactic Body Radiation Therapy in Patients With Hepatocellular Carcinoma. Front Oncol, 2019. 9: p. 896.

55. Bernard, V., et al., Single-Cell Transcriptomics of Pancreatic Cancer Precursors Demonstrates Epithelial and Microenvironmental Heterogeneity as an Early Event in Neoplastic Progression. Clin Cancer Res, 2019. 25(7): p. 2194–2205.

56. Meyer, M.A., et al., Breast and pancreatic cancer interrupt IRF8-dependent dendritic cell development to overcome immune surveillance. Nat Commun, 2018. 9(1): p. 1250.

57. Tjomsland, V., et al., The desmoplastic stroma plays an essential role in the accumulation and modulation of infiltrated immune cells in pancreatic adenocarcinoma. Clin Dev Immunol, 2011. 2011: p. 212810.

58. Laoui, D., et al., The tumour microenvironment harbours ontogenically distinct dendritic cell populations with opposing effects on tumour immunity. Nat Commun, 2016. 7: p. 13720.

59. Binnewies, M., et al., Unleashing Type-2 Dendritic Cells to Drive Protective Antitumor CD4(+) T Cell Immunity. Cell, 2019. 177(3): p. 556–571 e16.

60. Boucher, Y., et al., Bevacizumab improves tumor infiltration of mature dendritic cells and effector T-cells in triple-negative breast cancer patients. NPJ Precis Oncol, 2021. 5(1): p. 62.

61. Huang, Y., et al., Vascular normalizing doses of antiangiogenic treatment reprogram the immunosuppressive tumor microenvironment and enhance immunotherapy. Proc Natl Acad Sci U S A, 2012. 109(43): p. 17561–6.

62. Fukumura, D., et al., Enhancing cancer immunotherapy using antiangiogenics: opportunities and challenges. Nat Rev Clin Oncol, 2018. 15(5): p. 325–340.

63. Halai, K., et al., ICAM-2 facilitates luminal interactions between neutrophils and endothelial cells in vivo. J Cell Sci, 2014. 127(Pt 3): p. 620–9.

64. Zhao, Y., et al., Targeting Vascular Endothelial-Cadherin in Tumor-Associated Blood Vessels Promotes T-cell-Mediated Immunotherapy. Cancer Res, 2017. 77(16): p. 4434–4447.

65. Drobni, Z.D., et al., Renin-angiotensin-aldosterone system inhibitors and survival in patients with hypertension treated with immune checkpoint inhibitors. Eur J Cancer, 2022. 163: p. 108–118.

