## Supplemental_figures_Tables for "Addition of losartan to FOLFORINOX and chemoradiation downregulates pro-invasion and immunosuppression-associated genes in locally advanced pancreatic cancer"

**Supplementary Table 1.** Top 30 upregulated and 30 downregulated genes losartan+FFX+CRT vs untreated and FFX+CRT vs untreated

### Losartan+FFX+CRT vs untreated

| Gene | log2 Fold Change | Gene | log2 Fold Change |
|---|---|---|---|
| CXCL12 | 3.107106263 | PRAME | -2.740720505 |
| S100A12 | 3.066079227 | MS4A1 | -2.393884687 |
| PLA2G1B | 2.844783611 | CD19 | -2.269109818 |
| IL6R | 2.536735393 | TNFSF10 | -2.192247917 |
| CDKN1A | 2.357279706 | CEACAM1 | -2.038986236 |
| CD36 | 2.333143978 | CRP | -1.942508565 |
| NCAM1 | 2.216656032 | CEACAM6 | -1.931150488 |
| C7 | 2.090085519 | LTB | -1.874620654 |
| PTGDR2 | 2.082487988 | TNFRSF13C | -1.862462008 |
| EGR1 | 2.03652398 | MST1R | -1.795221663 |
| TNFRSF10C | 1.993801871 | CD22 | -1.749943938 |
| CFD | 1.930190442 | CARD11 | -1.645795579 |
| C1S | 1.870169584 | TIGIT | -1.627713485 |
| A2M | 1.82732473 | APOE | -1.579813684 |
| C1R | 1.79386208 | IFNL2 | -1.562969207 |
| CDH5 | 1.686105347 | C8G | -1.486078969 |
| MRC1 | 1.67680126 | IKBKE | -1.474634008 |
| MME | 1.666333457 | BAGE | -1.471209973 |
| CD209 | 1.661840481 | CXCL6 | -1.444271288 |
| IL6 | 1.628468226 | CD1A | -1.441887561 |
| S100A8 | 1.617490827 | LILRA4 | -1.411892363 |
| MCAM | 1.601468829 | HAMP | -1.387331053 |
| NFATC2 | 1.597515063 | BLK | -1.37950213 |
| CXCR2 | 1.5938512 | CARD9 | -1.353466064 |
| NFATC1 | 1.585587277 | CD79B | -1.350584906 |
| ALCAM | 1.583855159 | CD207 | -1.338332006 |
| CEBPB | 1.579985577 | SPP1 | -1.30377095 |
| CCL14 | 1.568506524 | CMKLR1 | -1.28675557 |
| ITGA5 | 1.553775982 | TNFRSF13B | -1.282425921 |
| C3 | 1.552484483 | TMEFF2 | -1.28171177 |

**FFX+CRT vs untreated**

| Gene | log2 Fold Change | Gene | log2 Fold Change |
|---|---|---|---|
| CXCL12 | 2.786385284 | CD19 | -1.59556708 |
| S100A12 | 2.39670815 | TNFSF10 | -1.588809816 |
| IL6 | 2.364961048 | PASD1 | -1.520372446 |
| CHIT1 | 2.341397779 | C8G | -1.504489408 |
| CD36 | 2.26777669 | CD1A | -1.502463848 |
| NCAM1 | 2.109342028 | CEACAM1 | -1.476956818 |
| CFD | 2.071863327 | BAGE | -1.371877113 |
| C7 | 2.009130033 | ISG15 | -1.346348912 |
| S100A8 | 1.99921441 | TNF | -1.340331809 |
| CCL2 | 1.898207953 | SPINK5 | -1.294335679 |
| CDKN1A | 1.811771092 | MST1R | -1.251119707 |
| HLA-DRA | 1.798049862 | IKBKE | -1.203617298 |
| CD6 | 1.755758576 | CD207 | -1.201928878 |
| TNFRSF10C | 1.746069298 | LILRA4 | -1.200529145 |
| A2M | 1.729697292 | IL3 | -1.195641387 |
| IL6R | 1.728099539 | IL22RA2 | -1.158804154 |
| CD209 | 1.688962235 | FOXJ1 | -1.154825736 |
| CCL11 | 1.649294657 | LTB | -1.147464958 |
| CCL21 | 1.58581805 | TNFRSF13C | -1.141840485 |
| C1S | 1.573429793 | MAGEA3 | -1.137454041 |
| EGR1 | 1.548193175 | DDX58 | -1.12531104 |
| CCL18 | 1.547490033 | IFNA7 | -1.111884386 |
| CR1 | 1.534196132 | IL24 | -1.101148838 |
| HLA-DPB1 | 1.532657244 | CARD9 | -1.100616041 |
| SAA1 | 1.529826861 | IGLL1 | -1.098223435 |
| MRC1 | 1.50147795 | IL17RB | -1.097305889 |
| C1R | 1.472634867 | IL1A | -1.092380557 |
| LY86 | 1.421245952 | ELANE | -1.088190128 |
| MCAM | 1.41836483 | IL13 | -1.07314633 |
| SELL | 1.386822499 | TPTE | -1.064776441 |

**Supplementary Table 2.** Gene Set Enrichment Analysis results of losartan+FFX+CRT vs FFX+CRT

| Gene Sets | NES | p-value | FDR q-value |
|---|---|---|---|
| GOMF DNA BINDING TRANSCRIPTION REPRESSOR ACTIVITY | 2.03667 | 0.0 | 0.0532 |
| GOBP RHYTHMIC PROCESS | 2.02573 | 0.0 | 0.0285 |
| GOCC CHROMOSOME | 1.95736 | 0.0 | 0.0699 |
| GOMF DNA BINDING TRANSCRIPTION FACTOR ACTIVITY | 1.92847 | 0.0 | 0.0769 |
| GOCC CHROMATIN | 1.90216 | 0.0 | 0.0860 |
| GOBP CIRCADIANRHYTHM | 1.886687 | 0.0019 | 0.0902 |
| GOMF SEQUENCE SPECIFIC DNA BINDING | 1.8675246 | 0.0 | 0.1027 |
| GOBP REGIONALIZATION | 1.8666339 | 0.0060 | 0.0908 |
| GOBP MAMMARY GLAND DEVELOPMENT | 1.8660303 | 0.0 | 0.0813 |
| GOMF TRANSCRIPTION FACTOR BINDING | 1.8650856 | 0.0 | 0.0744 |
| GOCC TRANSCRIPTION REGULATOR COMPLEX | 1.859656 | 0.0 | 0.0743 |
| GOMF CIS_REGULATORY REGION SEQUENCE SPECIFIC DNA BINDING | 1.849835 | 0.0 | 0.0776 |
| GOMF TRANSCRIPTION REGULATOR ACTIVITY | 1.8459487 | 0.0 | 0.0749 |
| GOBP CELL AGING | 1.830189 | 0.0 | 0.0866 |
| GOBP CHROMOSOME ORGANIZATION | 1.8002028 | 0.0041 | 0.1165 |
| GOBP RESPONSE TO UV | 1.7854171 | 0.0038 | 0.1301 |
| GOBP REGULATION OF MITOCHONDRION ORGANIZATION | 1.7661172 | 0.0065 | 0.1520 |
| GOBP MITOTIC CELL CYCLE PHASE TRANSITION | 1.7641045 | 0.0060 | 0.1467 |
| GOCC NUCLEOLUS | 1.7618381 | 0.0039 | 0.1428 |
| GOBP HEAD DEVELOPMENT | 1.7540505 | 0.0120 | 0.1483 |
| GOBP REGULATION OF AUTOPHAGY | 1.7434623 | 0.0060 | 0.1279 |
| GOBP RESPONSE TO RADIATION | 1.6620013 | 0.0099 | 0.1717 |

**Supplementary Table 3.** DEGs losartan+FFX+CRT vs untreated and FFX+CRT vs untreated

Losartan+FFX+CRT vs untreated

| Gene | baseMean | log2FoldChange | lfcSE | stat | p value | p adjusted |
|---|---|---|---|---|---|---|
| IFNG | 29.6420596 | -0.744415402 | 0.29152165 | -2.553551 | 0.01066307 | 0.02904492 |
| IL12A | 32.5976109 | -0.939543934 | 0.2645517 | -3.5514568 | 0.0003831 | 0.00190454 |
| IL4 | 20.7170759 | -0.927080453 | 0.34959378 | -2.6518791 | 0.00800452 | 0.0231877 |
| IL13 | 24.1710259 | -1.063508348 | 0.40282009 | -2.6401572 | 0.00828676 | 0.02391041 |
| IL23A | 34.9089588 | -0.874108434 | 0.29462679 | -2.9668329 | 0.00300884 | 0.0104097 |
| CD1A | 28.2896857 | -1.441887561 | 0.40173532 | -3.5891481 | 0.00033176 | 0.00170553 |

FFX+CRT vs untreated

| Gene | baseMean | log2FoldChange | lfcSE | stat | p value | p adjusted |
|---|---|---|---|---|---|---|
| IFNG | 29.6420596 | -0.758509704 | 0.28363786 | -2.6742189 | 0.00749036 | 0.02641526 |
| IL12A | 32.5976109 | -0.643382038 | 0.25519082 | -2.5211802 | 0.01169619 | 0.03811706 |
| IL4 | 20.7170759 | -0.941091626 | 0.34002277 | -2.7677312 | 0.0056448 | 0.021462 |
| IL13 | 24.1710259 | -1.07314633 | 0.39240174 | -2.7348154 | 0.00624153 | 0.02301169 |
| IL23A | 34.9089588 | -1.048044697 | 0.28768372 | -3.6430448 | 0.00026943 | 0.00202768 |
| CD1A | 28.2896857 | -1.502463848 | 0.39163159 | -3.8364215 | 0.00012484 | 0.00109799 |
| CCL21 | 697.045661 | 1.58581805 | 0.50631624 | 3.13207031 | 0.00173578 | 0.00892339 |

**Supplementary Table 4.** Genes included in panels B and C of Figure 3

**Dendritic cells**

CCR7

CD40LG

FT3LG

**TCR-CD3 complex**

ZAP70

CD3E

CD247

CD3D

LCK

ITK

**T cell activation & cytolytic activity**

CD5

SPN

NFATC4

IL16

GZMM

**Innate & adaptive immune response**

SH2D1A

SLAMF6

LY9

TLR9

**Inflammatory mediators**

LTB

IL11RA

CR2

**B cells**

BLK

MS4A1

CD79B

TNFRSF13B

TNFRSF13C

POU2F2

**Supplementary Table 5.** Genes included in panels C and D of Figure 4

**Tumor Suppressors**
RORA
CYLD
FEZ1
**Blood vessels**
NFATC4*
AKT3
PECAM1
CDH5
JAM3
**Dendritic cells**
IL3RA
FLT3LG
CSF1
**MHCII**
HLA-DRB3
LAMP2
**T cell activation**
STAT4
CD6
TNFSF8
**Inflammation Inhibition**
NCF4
SERPING1
NFKBIA
A2M
**Epithelial**
EPCAM
CDH1

**Supplementary Table 6.** Losartan+FFX+CRT correlation of individual genes with overall survival

| Positive correlation | | Negative correlation | |
|---|---|---|---|
| **Gene** | **Spearman correlation** | **Gene** | **Spearman correlation** |
| NFATC4 | 0.5 | GNLY | -0.67 |
| ENTPD1 | 0.5 | F12 | -0.6 |
| IL3RA | 0.5 | TTK | -0.59 |
| ELK1 | 0.5 | LYN | -0.59 |
| CDKN1A | 0.5 | SYK | -0.58 |
| CSF1 | 0.5 | BTLA | -0.56 |
| SERPING1 | 0.5 | ITGA6 | -0.56 |
| S100B | 0.5 | IL15RA | -0.55 |
| STAT3 | 0.6 | PBK | -0.54 |
| FOS | 0.6 | IL12RB2 | -0.54 |
| STAT2 | 0.6 | IDO1 | -0.54 |
| RORA | 0.6 | LGALS3 | -0.54 |
| GTF3C1 | 0.6 | CCR3 | -0.53 |
| THY1 | 0.6 | EPCAM | -0.53 |
| BCL2 | 0.6 | F2RL1 | -0.53 |
| PPBP | 0.6 | CASP1 | -0.52 |
| CFI | 0.6 | TNFSF15 | -0.51 |
| CD63 | 0.6 | CDH1 | -0.5 |
| EGR1 | 0.6 | | |
| MAPK11 | 0.6 | | |
| CD34 | 0.6 | | |
| CD99 | 0.6 | | |
| CD200 | 0.6 | | |
| TNFSF12 | 0.6 | | |
| AKT3 | 0.7 | | |
| ENG | 0.7 | | |
| UBC | 0.7 | | |
| PNMA1 | 0.7 | | |
| FEZ1 | 0.8 | | |
| JAM3 | 0.8 | | |

**Supplementary Table 7.** FFX+CRT correlation of individual genes with overall survival

| Positive correlation | | Negative correlation | |
|---|---|---|---|
| **Gene** | **Spearman correlation** | **Gene** | **Spearman correlation** |
| CXCL12 | 0.5 | TNFRSF4 | -0.75 |
| POU2AF1 | 0.5 | SERPINB2 | -0.65 |
| CD1C | 0.5 | HSD11B1 | -0.64 |
| BTLA | 0.5 | RIPK2 | -0.58 |
| PAX5 | 0.51 | ITGB1 | -0.57 |
| XCR1 | 0.51 | CXCL1 | -0.56 |
| CD209 | 0.52 | CD274 | -0.55 |
| BLNK | 0.52 | BID | -0.54 |
| PTGDR2 | 0.53 | TAP1 | -0.52 |
| POU2F2 | 0.57 | USP9Y | -0.51 |
| ECSIT | 0.57 | IRF1 | -0.5 |
| IRF4 | 0.57 | HLA-E | -0.49 |
| ADORA2A | 0.6 | BATF | -0.49 |
| FLT3 | 0.6 | | |
| TIRAP | 0.65 | | |
| CD1E | 0.68 | | |

**Supplementary Table 8:** Gene grouping association with overall survival

| Gene Sets | p value* |
|---|---|
| **Losartan+FFX+CRT: negative correlation** | |
| F12 TTK LYN SYK BTLA ITGA6 IL15RA PBK IDO1 LGALS3 CCR3 EPCAM F2RL1 CASP1 TNFSF15 CDH1 IL12RB GNLY | < 0.0001 |
| BTLA IDO1 LGALS3 EPCAM PBK ITGA6 TTK | 0.0006 |
| TTK PBK EPCAM ITGA6 | 0.0022 |
| IDO1 LGALS3 BTLA | 0.0042 |
| **Losartan+FFX+CRT: positive correlation** | |
| ENG CD200 JAM3 FEZ1 EGR1 RORA IL3RA CSF1 NFATC4 PNMA1 UBC AKT3 TNFSF12 CD99 CD34 MAPK11 CD63 CFI PPBP BCL2 THY1 GTF3C1 STAT2 STAT3 FOS S100B SERPING1 CDKN1A ELK1 ENTPD1 | 0.0538 |
| ENG JAM3 CD99 EGR1 RORA IL3RA CSF1 PNMA1 AKT3 MAPK11 GTF3C1 FOS CDKN1A ELK1 PPBP | 0.0138 |
| **FFX+CRT: negative correlation** | |
| TNFRSF4 SERPINB2 HSD11B1 RIPK2 ITGB1 CXCL1 CD274 BID TAP1 USP9Y IRF1 HLA-E BATF | 0.0417 |
| BID TAP1 USP9Y IRF1 HLA-E BATF HSD11B1 RIPK2 CXCL1 | 0.0043 |
| BID USP9Y BATF HSD11B1 | 0.0004 |
| **FFX+CRT: positive correlation** | |
| CD1E TIRAP FLT3 ADORA2A IRF4 ECSIT POU2F2 PTGDR2 BLNK PAX5 BTLA POU2AF1 CXCL12 CD1C CD209 XCR1 | 0.038 |
| CD209 CD1C FLT3 BTLA IRF4 CD1E ECSIT TIRAP CXCL12 | 0.0074 |
| CD209 CD1C FLT3 | 0.0014 |
| CD209 CD1C | 0.0006 |

*** Overall survival cutoff: 36 M**

**Supplementary Table 9.** Gene signature association with overall survival

| Gene Sets | p value* | p value** |
|---|---|---|
| CCL13 CCL17 CCL22 HSD11B1 CD209 | 0.7291 | 0.0523 |
| CCL13 CCL17 CCL22 CD209 | 0.6032 | 0.0013 |

*** Losartan+FFX+CRT OS cutoff 36 M**

**** FFX+CRT OS cutoff 36 M**

### Supplementary Figure 1

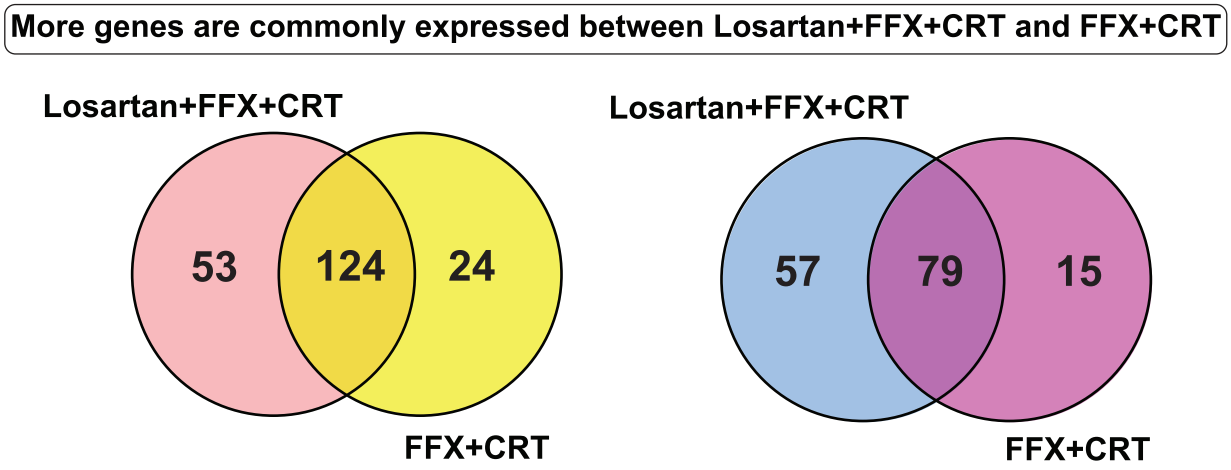

### Supplementary Figure 2

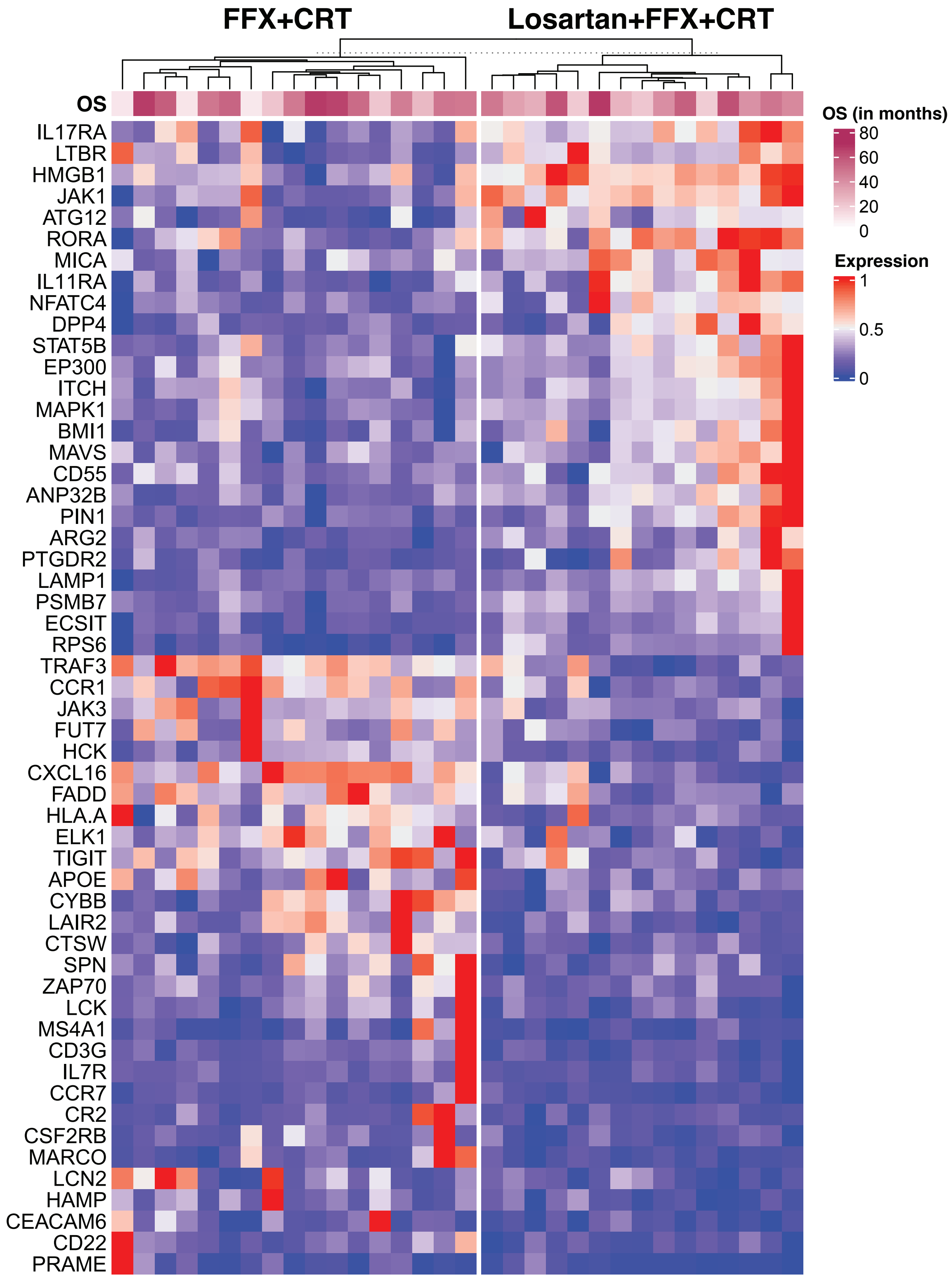

### Supplementary Figure 3

**A**

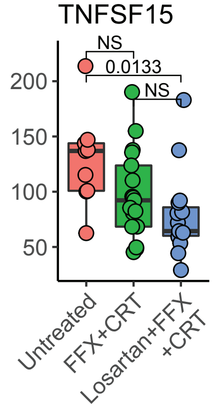

**B**

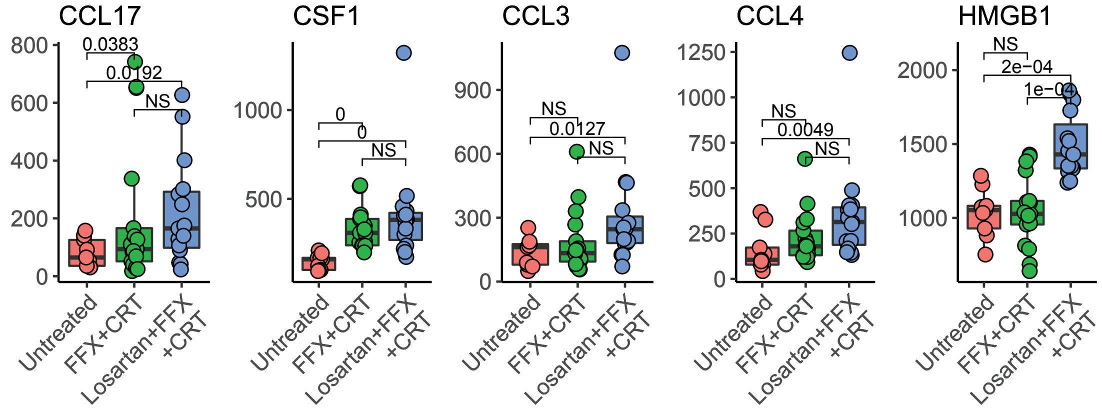

### Supplementary Figure 4

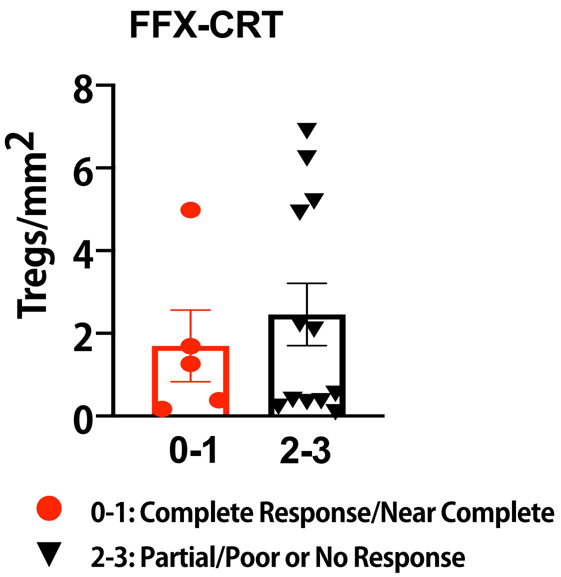
